# Safety, Tolerability, Pharmacokinetics, and Pharmacodynamics of Multiple Ascending Doses of mocravimod in Healthy Volunteers

**DOI:** 10.64898/2026.05.22.26353846

**Authors:** Dymphy Huntjens, Dirk Klingbiel, Jens Hasskarl

## Abstract

**Background:** Mocravimod is an oral sphingosine-1-phosphate (S1P) receptor modulator. This Phase 1 multiple-ascending-dose study evaluated its safety, tolerability, pharmacokinetics (PK), and pharmacodynamics (PD) in healthy volunteers.

**Methods:** In this double-blind, randomized, placebo-controlled, parallel-group trial, 60 healthy male volunteers were enrolled in five cohorts. Mocravimod was administered once daily at 0.3, 0.6, 1.2, or 3.0 mg for 14 days, or at 2.0 mg for 28 days. Safety assessments included adverse events (AEs), laboratory tests, vital signs, electrocardiography, and Holter monitoring. PK of mocravimod and its active metabolite, mocravimod-phosphate, and PD effects on absolute lymphocyte count (ALC) and leukocyte subsets were assessed.

**Results:** Fifty-nine of 60 participants completed the study. One participant in the 3.0 mg cohort discontinued treatment because of asymptomatic transaminase elevation. No deaths or serious AEs occurred. AEs were mostly mild or moderate, transient, and showed no clear dose relationship. Mocravimod produced dose-dependent reductions in ALC from 0.6 mg onward, with maximum geometric mean reductions of 65%, 74%, 83%, and 77% at 0.6, 1.2, 2.0, and 3.0 mg, respectively. ALC values recovered to above the lower limit of normal during follow-up in all cohorts. Holter monitoring showed an initial placebo-corrected reduction in heart rate of approximately 10–15 beats/min at doses of 1.2–3.0 mg, which attenuated with continued dosing. One participant in the 3.0 mg cohort had a recurrent daytime second-degree atrioventricular block (Mobitz I/Wenckebach), reported as a mild non-dose-limiting AE. No QT prolongation was observed. Exposure to mocravimod and mocravimod-phosphate increased approximately dose-proportionally. Steady state was reached by Day 14 (Day 28 in the 2.0 mg cohort), accumulation was approximately five- to sevenfold, terminal half-lives were approximately 100–140 hours for both analytes, and parent-to-metabolite exposure ratios were close to 1.

**Conclusions:** Once-daily mocravimod up to 3.0 mg for 14 days and 2.0 mg for 28 days was generally well tolerated and showed predictable S1P-modulator class effects on lymphocyte counts and heart rate, with PK properties supporting once-daily dosing and further clinical development.

## Introduction

Sphingosine 1-phosphate (S1P) receptor modulators have significantly advanced the treatment of immune-mediated disorders, including multiple sclerosis (MS), by selectively regulating lymphocyte trafficking. Fingolimod, the first agent in this class, established that functional antagonism of the S1P isoform 1 (S1P1) receptor results in lymphocyte sequestration within secondary lymphoid organs, thereby limiting their egress into the circulation and attenuating pathogenic immune activity [1]. This mechanism forms the basis for the clinical efficacy of S1P modulators in MS and has supported the development and regulatory approval of several second-generation compounds—siponimod, ozanimod, ponesimod, and etrasimod—for the treatment of MS and ulcerative colitis. These newer agents exhibit enhanced receptor selectivity and improved safety profiles compared with fingolimod [1–4]. S1P receptor modulators are being actively investigated across a broad spectrum of diseases, including neurodegenerative and inflammatory conditions, reflecting their capacity to modulate immune cell migration without inducing generalized immunosuppression [2, 5].

Mocravimod (KRP203) is an S1P modulator with strong affinity for the S1P1 and S1P5 receptors [1]. In contrast to conventional immunosuppressive therapies, S1P receptor modulators do not compromise T-cell cytotoxic function, thereby preserving anti-tumour immune responses [6]. Mocravimod is administered as a prodrug and undergoes *in vivo* phosphorylation to generate its active metabolite, mocravimod-phosphate.

Mocravimod is being investigated as adjunctive and maintenance therapy for acute myeloid leukaemia (AML) in the setting of allogeneic haematopoietic cell transplantation (allo-HCT) (NCT05429632). Its proposed therapeutic activity in the setting of allo-HCT is mediated by lymphocyte trafficking to lymphatic tissue and the bone marrow. The prevention of lymphocyte egress from the bone marrow compartment to the periphery results in a reduced absolute lymphocyte count, which, however, is not regarded as an adverse event as it is the main pharmacodynamic (PD) effect [7]. Mocravimod has demonstrated efficacy in preclinical models of autoimmune disease, transplantation, and haematological malignancies [7].

The present study investigated pharmacokinetic (PK), PD, and safety of multiple ascending dosing (MAD) of mocravimod when administered to healthy participants.

## Materials and Methods

### Study Design and Participants

This Phase 1, double-blind, randomized, placebo-controlled, parallel-group MAD study (EudraCT 2007-005608-42) was conducted at two sites in the United Kingdom. Sixty healthy adults (male; 18–55 years; non-smokers; BMI 18–30 kg/m²) were randomized in 5 cohorts (n=12 each; 9 active/3 placebo). Cohorts 1–3 and 5 received once-daily mocravimod at 0.3, 0.6, 1.2, and 3.0 mg for 14 days; Cohort 4 received 2.0 mg for 28 days. In-patient monitoring occurred through Day 16 (cohorts 1–3,5) or Day 30 (cohort 4) and then followed to Day 35 or Day 49, respectively (Figure 1). The dose range was selected based on the active dose range in the single dose healthy volunteer trial [8]. The starting dose 0.3 mg was selected to have minimal initial effects in terms of reduced blood lymphocyte counts or heart rates.

**Figure 1.**
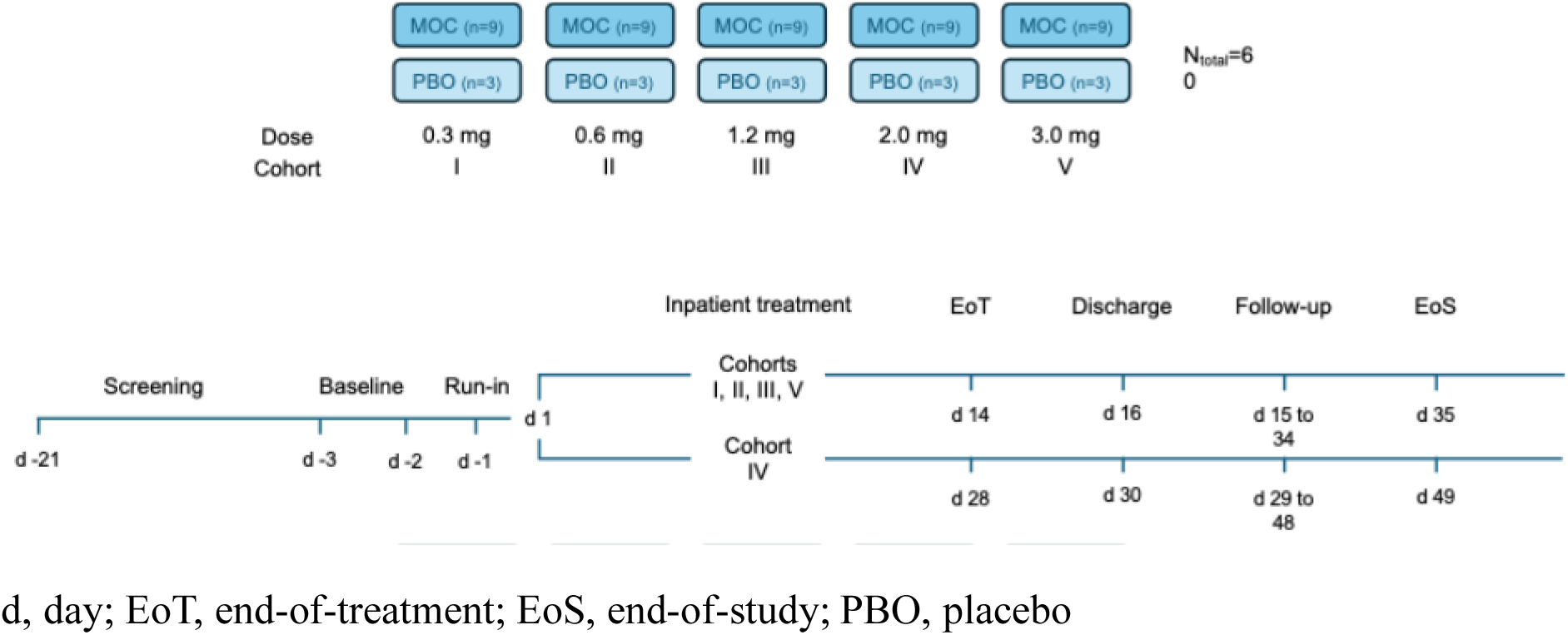
Study schematic. d, day; EoT, end-of-treatment; EoS, end-of-study; PBO, placebo

### Ethics

The study was designed, implemented, and reported in accordance with the International Conference on Harmonization (ICH) Harmonized Tripartite Guidelines for Good Clinical Practice (GCP), and with the ethical principles laid down in the Declaration of Helsinki. Informed consent was obtained from each participant in writing before randomization.

### Randomization and Blinding

Randomization employed ascending (Site 1) and descending (Site 2) sequences per cohort; placebo was identical in appearance to active drug. Pharmacists and bioanalysts were unblinded for preparation/analysis; investigators and participants remained blinded. Emergency code breaks were available but unused.

### Treatments and Dosing

Mocravimod capsules (0.1 mg and 1 mg strengths) and matching placebo were administered once daily between 08:30–09:30 with 180–240 mL water. Participants fasted for approx. 10 hours prior to and one hour post-dose with no food or fluid intake apart from the fluid given at the time of drug intake. No dose adjustments or planned interruptions were permitted.

### Safety and Tolerability Assessments

Safety monitoring encompassed adverse and severe adverse events (AEs/SAEs), clinical labs (haematology, chemistry, urinalysis), vital signs (supine/standing), 12-lead ECGs, 24-hours Holter monitoring, ambulatory blood pressure monitoring (ABPM) (Days −1, 8, 14; and Day 28 for cohort 4), pulmonary function tests (PFTs; spirometry, forced expiratory volume in one second [FEV1], forced expiratory flow at 25–75% of forced vital capacity [FEF25–75%]), ophthalmologic exams and optical coherence tomography (OCT), Borg breathlessness scale, neurologic examinations (including mini-mental status examination [MMSE]), and body weight. Special safety biomarkers (urinary kidney injury molecule 1 [KIM-1], neutrophil gelatinase-associated lipocalin [NGAL], cystatin C; blood alpha-glutathione S-transferase [α-GST]) were explored.

### Pharmacokinetics

Whole blood PK sampling for mocravimod and mocravimod-phosphate was performed at Day 1 at the following time points: pre-dose, and post-dose at 0.25, 0.5, 1, 2, 3, 4, 6, 8, 10, 12, 16 hours post dose. On Day 2, 4, 8 and 12 a pre-dose sample was collected. On Day 14, at the following time-points: pre-dose, 0.25, 0.5, 1, 2, 3, 4, 6, 8, 10, 12, 16, and 24 (Day 15), 36, 48 (Day 16), 96 (Day 18) hours post-dose (Cohort 1, 2, 3 and 5). On Days 21, 24 and 28, a sample was taken at a similar time to the intake on Day 14. For the 2.0 mg cohort 4, additional sampling was performed on Day 28 pre-dose, 0.5, 1, 2, 4, 6, 8, 12, 24 (Day 29), 36, 48 (Day 30), 96 (Day 32) hours post-dose. On Days 35, 38 and 42, a sample was taken at a similar time to the intake on Day 28. A total of 35 samples per individual was collected for cohorts 1, 2, 3 and 5, whereas 51 samples were collected for cohort 4.

Urine samples were collected over 24-hour intervals on Days 1 and 14 (and Day 28 for Cohort 4), following procedures that included pooling urine during each interval, measuring total volume, and storing aliquots at –70°C for analysis. A validated LC-MS/MS assay (LLOQ 0.1 ng/mL for blood, LLOQ 1.5 ng/mL for urine) for both analytes was used. Non-compartmental methods were used to determine PK parameters such as AUC_last_, AUC_tau_, C_max_, C_min_ for both analytes.

### Pharmacodynamics

Samples for absolute lymphocyte count (ALC) assessments were collected on Day 1 pre-dose, 1, 2, 3, 4, 6, 8, 10, 12 hrs post-dose; pre-dose on Day 2, 3, 4, 5, 7, 10, 14, and in the morning of Days 21, 24, 28 and study completion (Day 35) (not 2 mg) and Day 38, 42 and 49. PD endpoints included ALC (absolute counts) at dense time points on Day 1 and across treatment/follow-up and flow-cytometric leukocyte subsets (CD3+, CD4+, CD8+, CD19+, NK, naïve/memory phenotypes, dendritic cells). Statistical models estimated geometric mean fractions from baseline, E_max_, time to minimum ALC, and recovery. Subsets were collected at baseline (Day −2), Days 14, 28, 42, and 49 (cohort 4) and baseline, Days 14, 28, and 35 (cohorts 1, 2, 3, 5).

## Results

### Participants and Disposition

The overall median age of participants in the safety analysis set was 29 years with a range of 19–52 years. The overall median height of participants in the safety analysis set was 177.5 cm, with a range of 159–199 cm. The overall median weight of participants in the safety analysis set was 80.15 kg, with a range of 60.9–115.7 kg. With respect to race, 59 participants (98.3%) were Caucasian, and 1 participant (1.7%) was classified as other, with a reported ethnicity of mixed race (African/Caucasian). All 60 (100%) participants were male. Across the active dose and placebo cohorts, the mean and median age, weight and body mass index (BMI) were comparable (Table 1).

**Table 1.** Demographics per Cohort.

|  | **Mocravimod** |  |  | **Placebo** |  | **Overall** |  |
| --- | --- | --- | --- | --- | --- | --- | --- |
|  | Cohort 1<br>0.3 mg<br>(N = 9)<br>n (%) | Cohort 2<br>0.6 mg<br>(N = 9)<br>n (%) | Cohort 3<br>1.2 mg<br>(N = 9)<br>n (%) | Cohort 4<br>2 mg<br>(N = 9)<br>n (%) | Cohort 5<br>3 mg<br>(N = 9)<br>n (%) | Pooled<br>Placebo<br>(N = 15)<br>n (%) | Overall<br>(N = 60)<br>n (%) |
| **Age (years)** |  |  |  |  |  |  |  |
| Mean (SD) | 26.3 (4.3) | 30.8 (8.3) | 36.6 (11.3) | 33.3 (8.9) | 34.7 (9.4) | 29.7 (9.0) | 31.7 (9.1) |
| Median | 25.0 | 29.0 | 35.0 | 34.0 | 31.0 | 27.0 | 29.0 |
| Range | 22-35 | 19-48 | 23-52 | 20-46 | 25-51 | 21-49 | 19-52 |
| **Gender, n (%)** |  |  |  |  |  |  |  |
| Male | 9 (100%) | 9 (100%) | 9 (100%) | 9 (100%) | 9 (100%) | 15 (100%) | 60 (100%) |
| **Race, n (%)** |  |  |  |  |  |  |  |
| Caucasian | 9 (100%) | 8 (89%) | 9 (100%) | 9 (100%) | 9 (100%) | 15 (100%) | 59 (98%) |
| Other |  | 1 (11%) |  |  |  |  | 1 (2%) |
| **Weight, (kg)** |  |  |  |  |  |  |  |
| Mean (SD) | 82.4 (9.1) | 78.6 (5.8) | 79.8 (7.8) | 82.7 (11.5) | 82.0 (8.2) | 80.6 (13.0) | 80.9 (9.6) |
| Median | 81.1 | 76.0 | 81.8 | 78.3 | 80.5 | 78.4 | 80.2 |
| Range | 69.5-102.8 | 71.4-88.0 | 69.0-90.7 | 65.2-102.5 | 71.5-93.7 | 60.9-115.7 | 60.9-115.7 |
| **Height, (cm)** |  |  |  |  |  |  |  |
| Mean (SD) | 180.6 (7.5) | 180.6 (6.9) | 175.9 (5.4) | 175.4 (7.1) | 174.0 (3.7) | 177.7 (9.3) | 177.4 (7.3) |
| Median | 180.0 | 180.0 | 178.0 | 175.0 | 175.0 | 178.0 | 177.5 |
| Range | 169-195 | 169-193 | 166-182 | 167-186 | 166-177 | 159-199 | 159-199 |
| **BMI (kg/m<sup>2</sup>)** |  |  |  |  |  |  |  |
| Mean (SD) | 25.2 (1.2) | 24.2 (2.5) | 25.8 (2.3) | 26.8 (2.8) | 27.1 (2.5) | 25.5 (3.1) | 25.7 (2.6) |
| Median | 25.3 | 23.0 | 25.7 | 27.6 | 27.4 | 26.4 | 26.2 |
| Range | 23.0-27.0 | 20.4-27.8 | 21.6-29.3 | 20.8-29.6 | 23.6-30.0 | 19.0-29.2 | 19.0-30.0 |

Abbreviations: BMI = body mass index; N = total number of participants in safety population; n = number of participants; SD = standard deviation

### Pharmacokinetics

Mean blood concentration-time profiles of mocravimod and mocravimod-phosphate on Day 1 and Day 14 or Day 28 are displayed in Figure 2. On day 1, mocravimod and mocravimod-phosphate blood concentrations decreased very slowly, following a mono-exponential pattern. Blood concentrations of both analytes appeared to increase proportionally to the dose. Summary statistics of blood concentrations of mocravimod and mocravimod-phosphate are presented in Table 2. Mocravimod appeared after a short lag (median Tlag 0.5–1.0 h), with median Tmax 2–6 h. Mocravimod-phosphate appeared in blood after some lag-time, longer than that of mocravimod yet more variable, ranging from 15 minutes to 4 hours. Tmax of mocravimod-phosphate was later compared to mocravimod parent (median Tmax ∼10 h). Exposure parameters C_max_ and AUC_last_ increased nearly proportionally over 0.6–3.0 mg (no reliable parameters at 0.3 mg). The metabolite/parent exposures were of similar magnitude (molar ratios were approximately 1).

**Figure 2.**
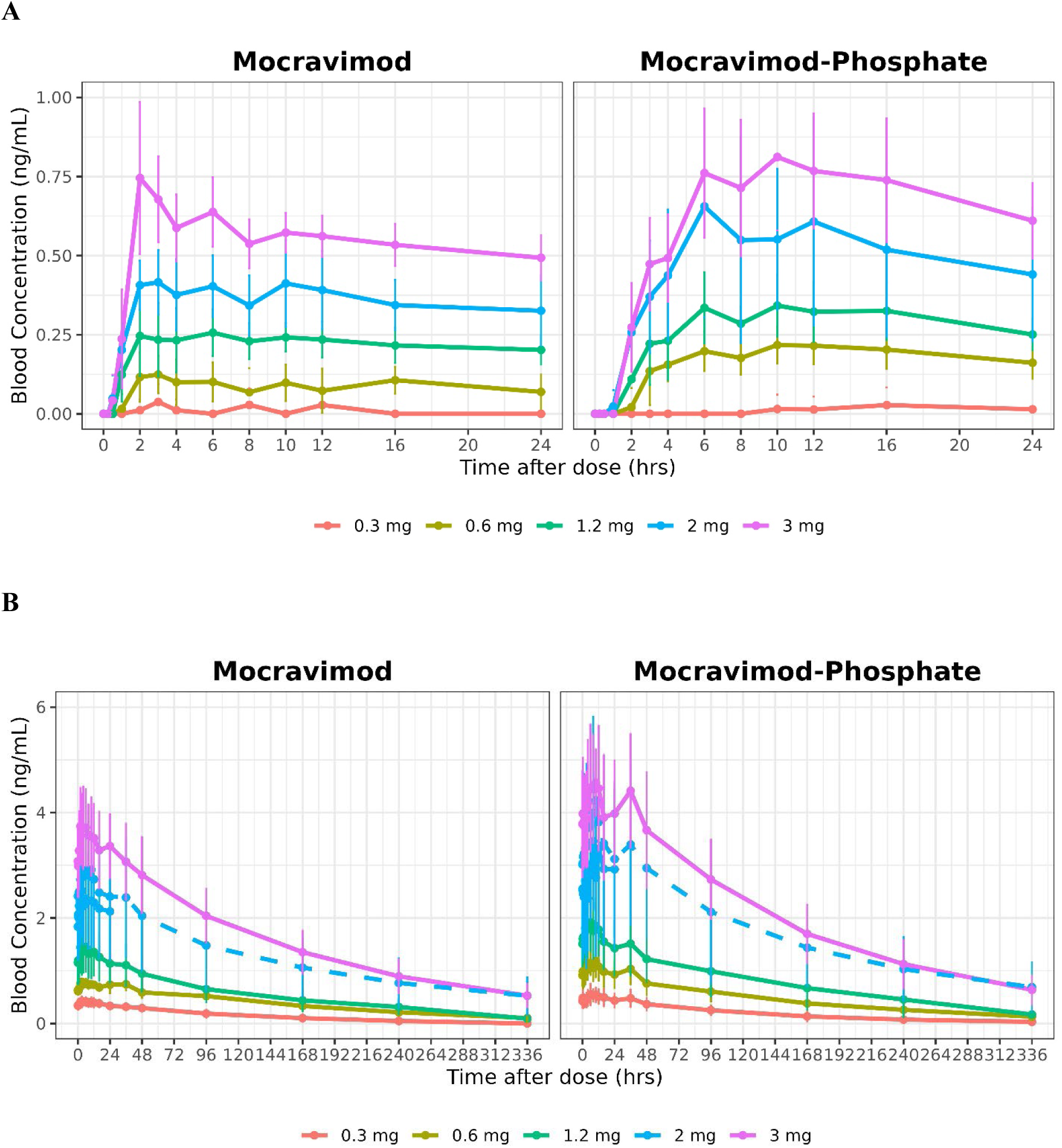
Mean blood concentration versus time by cohort (Day 1 (A) and Day 14 or Day 28 (B)). Day 1 Blood concentrations (ng/mL) of mocravimod and mocravimod-phosphate following 0.3, 0.6, 1.2, 2 and 3 mg mocravimod for 14 days (or 28 days for 2 mg arm) Solid lines indicate Day 14, and dashed lines indicate Day 28.

**Table 2:** Geometric mean blood pharmacokinetic parameter summary statistics by analyte, dose and by day.

|  | 0.3 mg |  | 0.6 mg |  | 1.2 mg |  | 2 mg |  |  | 3 mg |  |
| --- | --- | --- | --- | --- | --- | --- | --- | --- | --- | --- | --- |
| PK parameters | Day 1 | Day 14 | Day 1 | Day 14 | Day 1 | Day 14 | Day 1 | Day 14 | Day 28 | Day 1 | Day 14 |
| **Mocravimod** |  |  |  |  |  |  |  |  |  |  |  |
| n | 0 | 9 | 6 | 9 | 9 | 9 | 9 | 9 | 9 | 9 | 8 |
| C<sub>max</sub> (ng/mL) | NC | 0.433 (23) | 0.156 (21) | 0.834 (13) | 0.284 (33) | 1.43 (40) | 0.437 (26) | 2.38 (33) | 3.07 (45) | 0.804 (18) | 3.87 (20) |
| AUC<sub>last</sub> (h.ng/mL) | NC | 36.1 (61) | 2.45 (33) | 119 (23) | 4.54 (41) | 153 (62) | 8.31 (28) | 50.5 (36) | 374 (55) | 12.8 (14) | 527 (31) |
| AUC<sub>tau</sub> (h.ng/mL) |  | 9.0 (23) |  | 17.3 (12) |  | 28.8 (44) |  | 46.9 (29) | 58.9 (38) |  | 80.9 (21) |
| T<sub>max</sub> (h) | NC | 4.0 (2.0-10.0) | 2.5 (2.0-4.0) | 3.0 (2.30-24.0) | 6.0 (2.0-10.0) | 6.0 (0.25-10.0) | 3.0 (2.0-10.0) | 6.0 (2.0-16.0) | 6.0 (1.0-10.0) | 2.0 (2.0-6.0) | 3.0 (2.0-8.0) |
| T<sub>lag</sub> (h) | NC |  | 1.0 (0.5-1.0) |  | 0.5 (0.25-4.0) |  | 0.5 (0.25-0.5) |  |  | 0.5 (0.25-1.0) |  |
| T<sub>1/2</sub> (h) |  | NC |  | 117 (29) |  | 102 (21) |  | NC | 139 (31) | 121 (20) |  |
| **Mocravimod-phosphate** |  |  |  |  |  |  |  |  |  |  |  |
| n | 0 | 9 | 9 | 9 | 9 | 9 | 9 | 9 | 9 | 9 | 8 |
| C<sub>max</sub> (ng/mL) | NC | 0.612 (30) | 0.226 (25) | 1.21 (26) | 0.381 (29) | 2.06 (31) | 0.639 (53) | 3.41 (46) | 4.17 (43) | 0.884 (22) | 5.05 (22) |
| AUC<sub>last</sub> (h.ng/mL) | NC | 50 (67) | 3.95 (27) | 143 (38) | 6.01 (30) | 219 (65) | 10.6 (48) | 65.2 (47) | 525 (51) | 14.9 (22) | 675 (34) |
| AUC<sub>tau</sub> (h.ng/mL) |  | 11.3 (29) |  | 24.0 (27) |  | 38.4 (32) |  | 65.3 (47) | 78.9 (42) |  | 98.3 (25) |
| T<sub>max</sub> (h) | NC | 8.0 (6.0-16.0) | 10.0 (3.0-12.0) | 8.1 (2.0-10.1) | 10.0 (6.0-16.0) | 6.0 (1.0-12.0) | 10.0 (4.0-12.0) | 10.0 (8.0-12.0) | 10.0 (1.0-10.0) | 10.0 (6.0-16.0) | 10.0 (6.0-12.0) |
| T<sub>lag</sub> (h) | NC |  | 2.0 (1.0-3.0) |  | 1.0 (1.0-4.0) |  | 0.5 (0.25-1.0) |  |  | 1.0 (0.25-2.0) |  |
| T<sub>1/2</sub> (h) |  | NC |  | 121 (34) |  | 106 (34) |  | NC | 127 (32) | 113 (21) |  |
| C<sub>max</sub> Ratio<sup>1</sup> | NC | 1.20 (21) | 1.08 (31) | 1.23 (28) | 1.14 (31) | 1.22 (24) | 1.24 (43) | 1.21 (26) | 1.15 (40) | 0.932 (21) | 1.11 (20) |
| AUC Ratio<sup>1</sup> | NC | 1.07 (21) | 1.08 (31) | 1.17 (26) | 0.98 (26) | 1.13 (21) | 1.10 (33) | 1.09 (27) | 1.13 (32) | 0.97 (21) | 1.03 (17) |

Geometric mean (CV%) reported except for T<sub>max</sub> and T<sub>lag</sub>, for which median (min,max) values are reported. C<sub>max</sub> Ratio and AUC ratio are calculated based on molecular weight of mocravimod-phosphate versus mocravimod. 1) N=6 for C<sub>max</sub> ratio at dose 0.6 mg and AUC ratio at dose 1.2 mg. N=5 for AUC ratio at dose 0.6 mg. N=8 for AUC ratio at dose 2 and 3 mg.

Steady-state levels were achieved by Day 14 (Day 28 for 2.0 mg), with accumulation ratios ∼5–7 for both C_max_ and AUC. At steady-state, mocravimod C_max_, and AUC_tau_ increased approximately dose-proportionally (0.3–3.0 mg), the fluctuation indices were modest (≈25–46%), and terminal half-lives were long (≈100–140 h) for mocravimod and mocravimod-phosphate. Parent mocravimod was quantifiable in urine in some participants at ≥2 mg whereas mocravimod-phosphate was not quantifiable. These findings suggest minimal renal elimination of unchanged drug and negligible urinary excretion of the phosphate metabolite.

### Pharmacodynamics

The PD effects of mocravimod were determined by the changes in ALC over time, expressed in fraction from Day 1 pre-dose. On Day 1 ALC were collected pre-dose, 1, 2, 3, 4, 6, 8, 10, 12 and 24 hours post-dose. The Day 1 level in the mean ALC for all mocravimod dosing groups, except 0.3 mg, start to decrease compared to the placebo group around 3-4 hours post-dose and continues to stay below the placebo group until the latest observation 12 h post-dose. Visual comparison of the data in Figure 3A and B indicates a dose dependent reduction of mean ALC for all doses of mocravimod ≥0.6 mg. For the lowest dose, 0.3 mg, no change was noted on Day 1 when compared to placebo. With continued dosing, pre-dose ALC reductions were dose-dependent across 0.6–3.0 mg. Maximal reduction in ALC increased with dose (Table 3), with the maximal decrease (estimated mean of 83% reduction in ALC) observed in the 2.0 mg mocravimod dose group (28 days of treatment). No further reduction in ALC was observed at the higher (3.0 mg) dose group (14 days of treatment). Interestingly, the variability in the maximal observed lymphocyte effect appears reduced when the effect is large (such as for the 2 mg dose level). Gradual normalization of ALC (≥1.0 x10^9^/L) after 14 (or 28) days of daily mocravimod dosing was observed during the follow-up period. On Day 28, all dose groups except 0.3 mg showed still dose-dependent statistically significant decreases compared to placebo. Full return (100% of treated participants) to Day 1 pre-dose levels was observed by Day 35 only for the dose groups 0.3 and 0.6 mg (Table 3). However, a clear trend towards returning to Day 1 pre-dose levels was apparent for all dose groups and mean ALC levels were above the lower limit of normal (LLN; ≥1.0 x10^9^/L) by Day 35 (Day 49 for the 2 mg cohort). All individuals with ALC < LLN at Day 35 (Day 49 for the 2 mg cohort) were followed until ALCs were normal.

**Figure 3:**
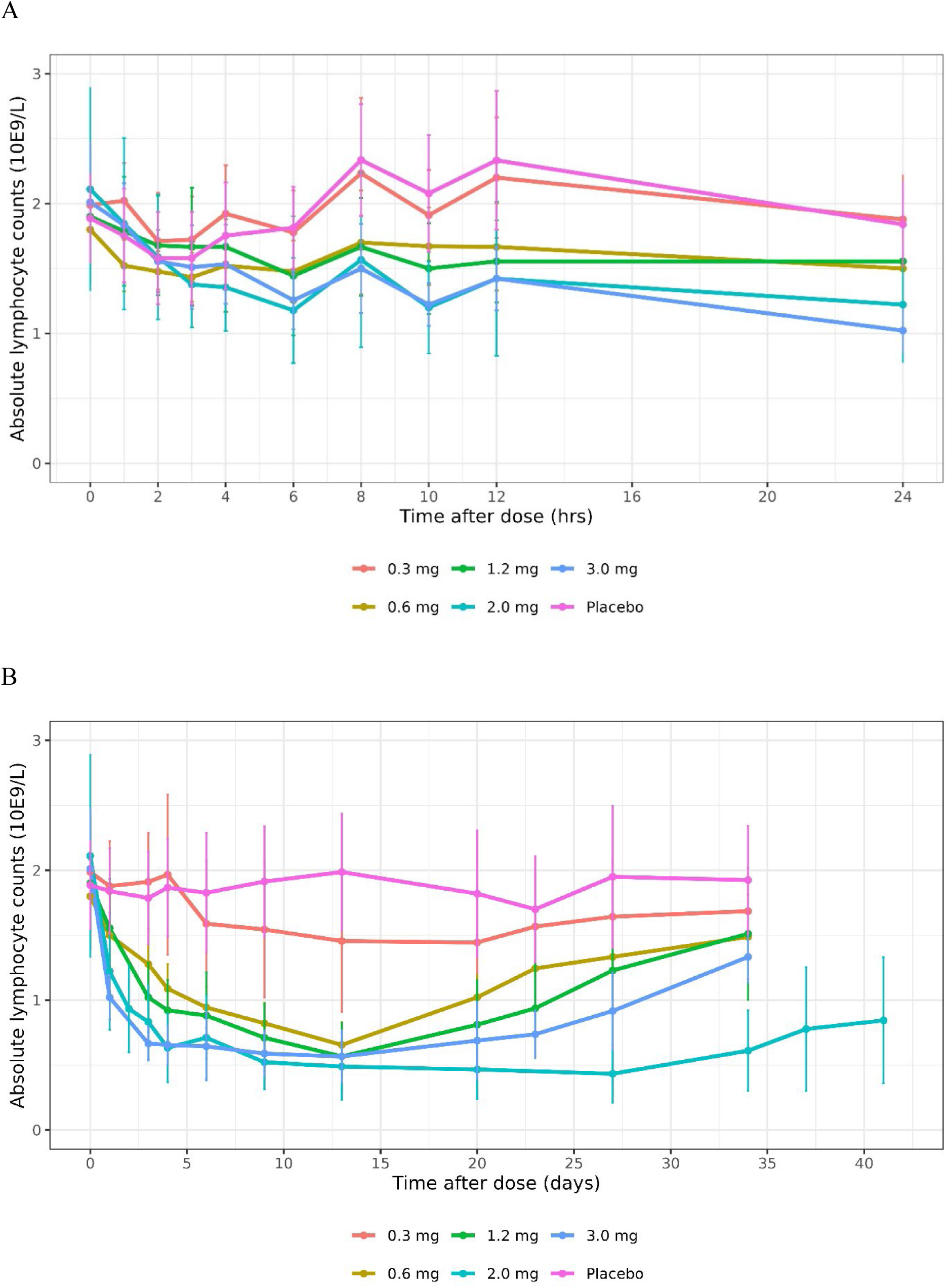
Absolute lymphocyte counts over time by dose group. Plot A shows first 24 hours after first dose intake and plot B shows the time course after multiple dosing in days until end of study. Results are shown and mean ± standard deviation.

**Table 3:** Pharmacodynamic results of lymphocyte counts.

| Treatment | GM Emax (%) | 95% CI | Emax (10E9/L) | Time to Emax (days) | Time to recovery (days) | Time to return to 1 10E9/L (days) |
| --- | --- | --- | --- | --- | --- | --- |
| Placebo | 29 | (18-39) | 0.725 (17) | 0.1 (0.0-48.0) | 6.0 (0.2-48.0) | 34.0 (20.0-48.0) |
| 0.3 mg | 39 | (26-49) | 0.625 (25) | 6.0 (0.1-23.0) | 34.0 (0.3-34.0) | 34.0 (13.0-34.0) |
| 0.6 mg | 65 | (58-71) | 0.361 (27) | 12.9 (8.9-13.0) | 34.0 (32.3-34.1) | 34.0 (20.0-34.1) |
| 1.2 mg | 74 | (68-78) | 0.279 (37) | 13.0 (8.9-20.0) | 34.1 (34.0-50.1) | 34.0 (23.0-34.1) |
| 2 mg | 83 | (80-86) | 0.174 (28) | 19.9 (9.0-37.0) | 48.0 (47.0-56.1) | 48.0 (47.0-56.1) |
| 3 mg | 77 | (72-81) | 0.243 (41) | 9.0 (3.0-20.0) | 34.0 (20.0-35.0) | 34.0 (20.0-35.0) |

GM= geometric mean, CI=confidence interval, GM E<sub>max</sub> is the maximum geometric mean percentage decrease in absolute lymphocyte counts from baseline. Mean (CV%) reported except for Time to E<sub>max</sub>, Time to recovery and time to return to 1 10E9/L, for which median (min,max) values are reported

Relative levels of both T cells (CD3+) and B cells (CD19+) were reduced in a dose dependent manner in the mocravimod 0.6 mg to 3 mg dose groups on Day 14 of the treatment administration phase as compared to baseline, while the mean values of the placebo group remained stable (Figure 4). The two major T lymphocyte subpopulations, CD4+ and CD8+ cells, were affected to different degrees by mocravimod (Figure 4). While CD4+ cells showed a clear dose-dependent reduction among all T lymphocytes (Day 14 mean ratios versus placebo ranging from 0.65 [95% CI: 0.51–0.84] in the mocravimod 0.6 mg dose group to 0.29 [95% CI: 0.22–0.37] in the mocravimod 3.0 mg dose group), the relative proportion of CD8+ T cells was found to increase. The most pronounced increase was observed for mocravimod 2.0 mg, on Days 14 (geometric mean ratio of 1.93 [95% CI: 1.65–2.25] and 28 (geometric mean ratio of 1.93 [95% CI: 1.66–2.25]), when compared to the placebo group.

**Figure 4:**
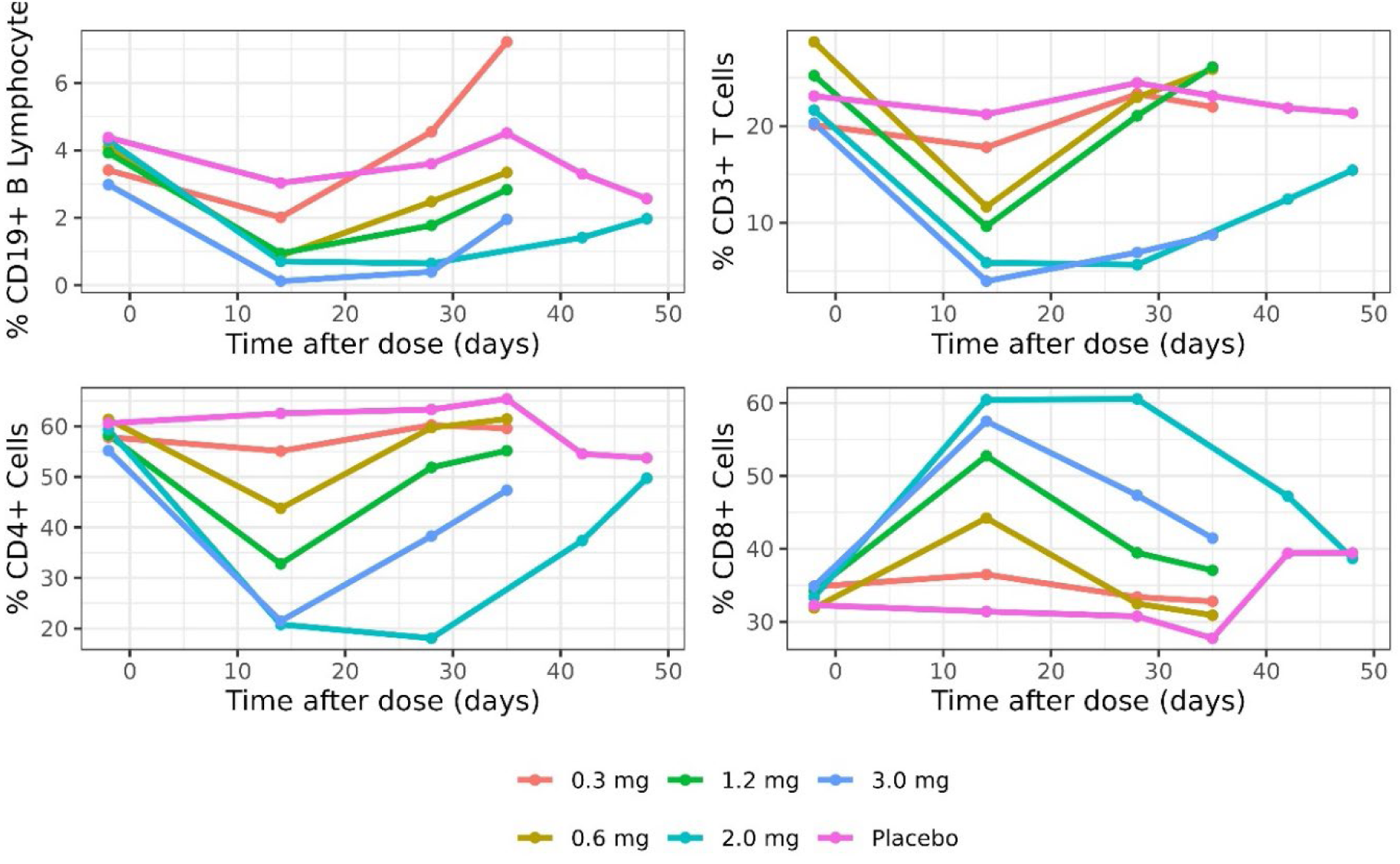
Mean CD3+ T cells and CD19+ B lymphocytes as percentage of leukocytes, CD4+ and CD8+ cells as percentage of CD3+ T cells by dose group.

### Safety and Tolerability

AEs were assessed from the time of first administration of study drug until study completion. No deaths or SAEs occurred. A total of 155 AEs were reported in 45 participants (75%) across active and placebo arms and were largely mild/moderate and transient; their frequency was similar across doses and placebo. The most frequently affected system organ classes (SOC), which involved ≥5% of participants, were nervous system disorders (21 participants, 35%), respiratory, thoracic and mediastinal disorders (15 participants, 25%). general disorders and administration site conditions (11 participants, 18.3%), gastrointestinal disorders (10 participants, 16.7%), investigations (9 participants, 15%), musculoskeletal and connective tissue disorders (8 participants, 13.3%), infections and infestations (7 participants, 11.7%), psychiatric disorders (5 participants, 8.3%), and skin and subcutaneous tissue disorders (3 participants, 5%). The AEs reported by ≥5% of participants were nasopharyngitis (4 participants, 6.7%), cough (3 participants, 5%), dyspnoea (3 participants, 5%), nasal congestion (4 participants, 6.7%), orophayngeal pain (6 participants, 10%), rhinorrhoea (3 participants, 5%), headache (15 participants, 25%), nausea (3 participants, 5%), dizziness (5 participants, 8.3%), back pain (4 participants, 6.7%), ALT increased (6 participants, 10%) and AST increased (5 participants, 8.3%). There appeared to be no important relationship between dose and frequency of AEs (Table 4).

**Table 4A:**
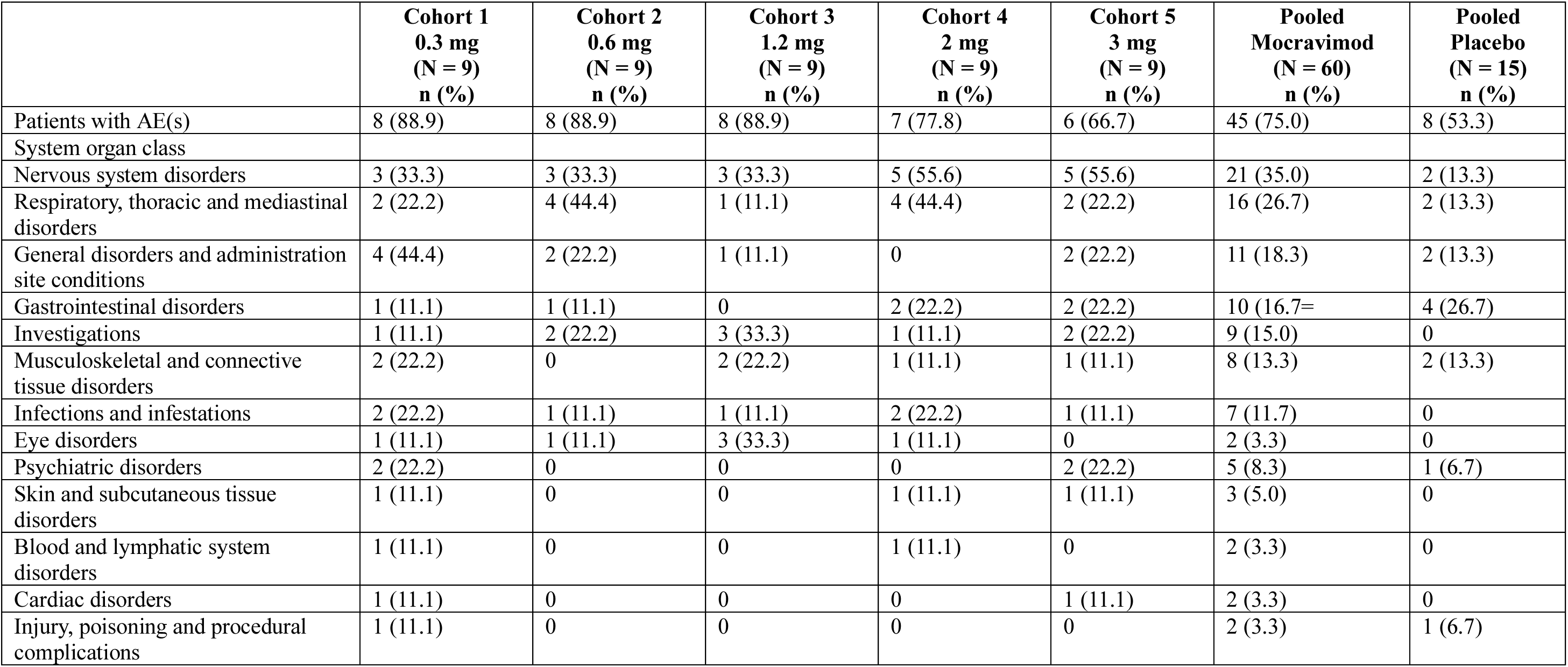
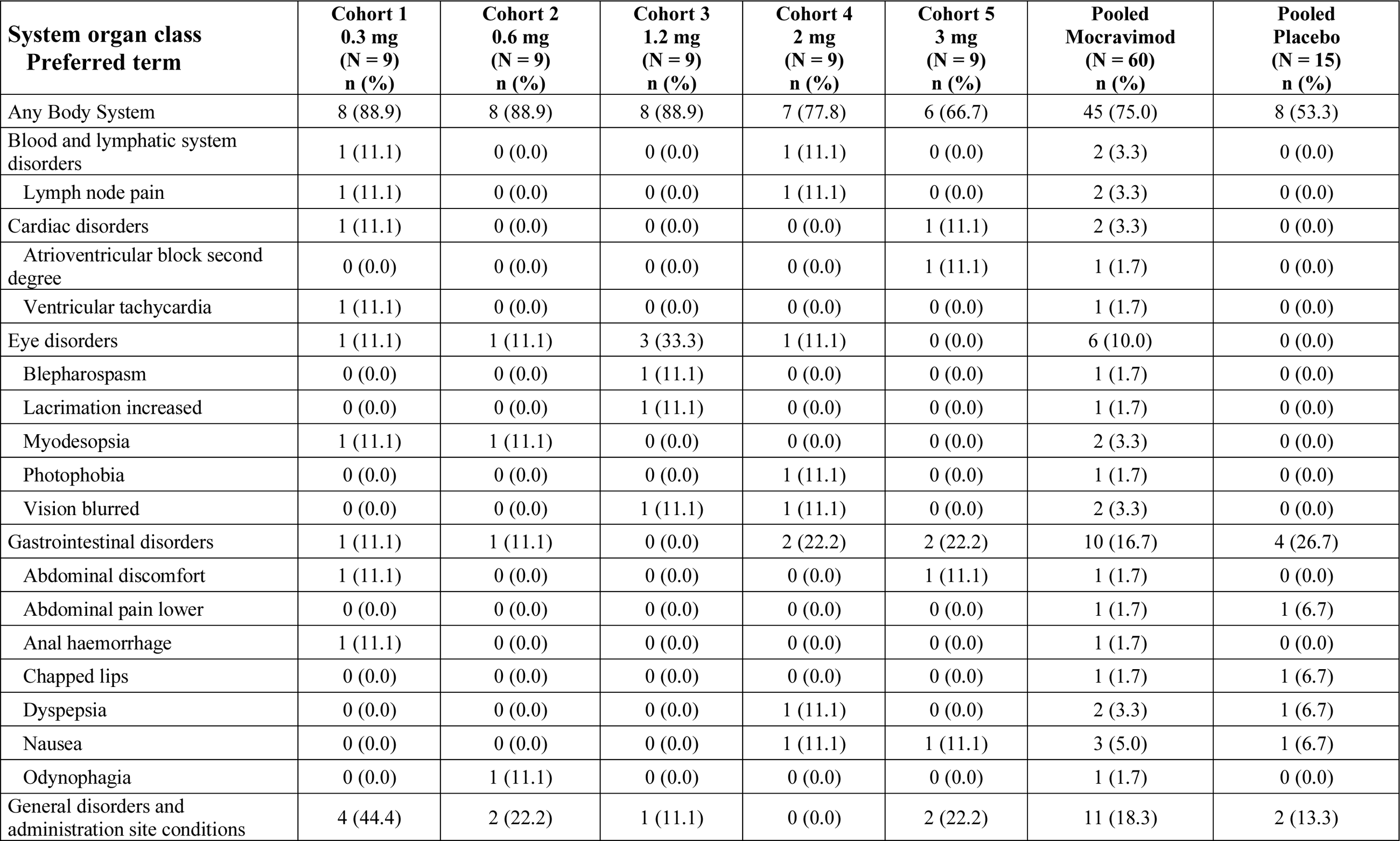
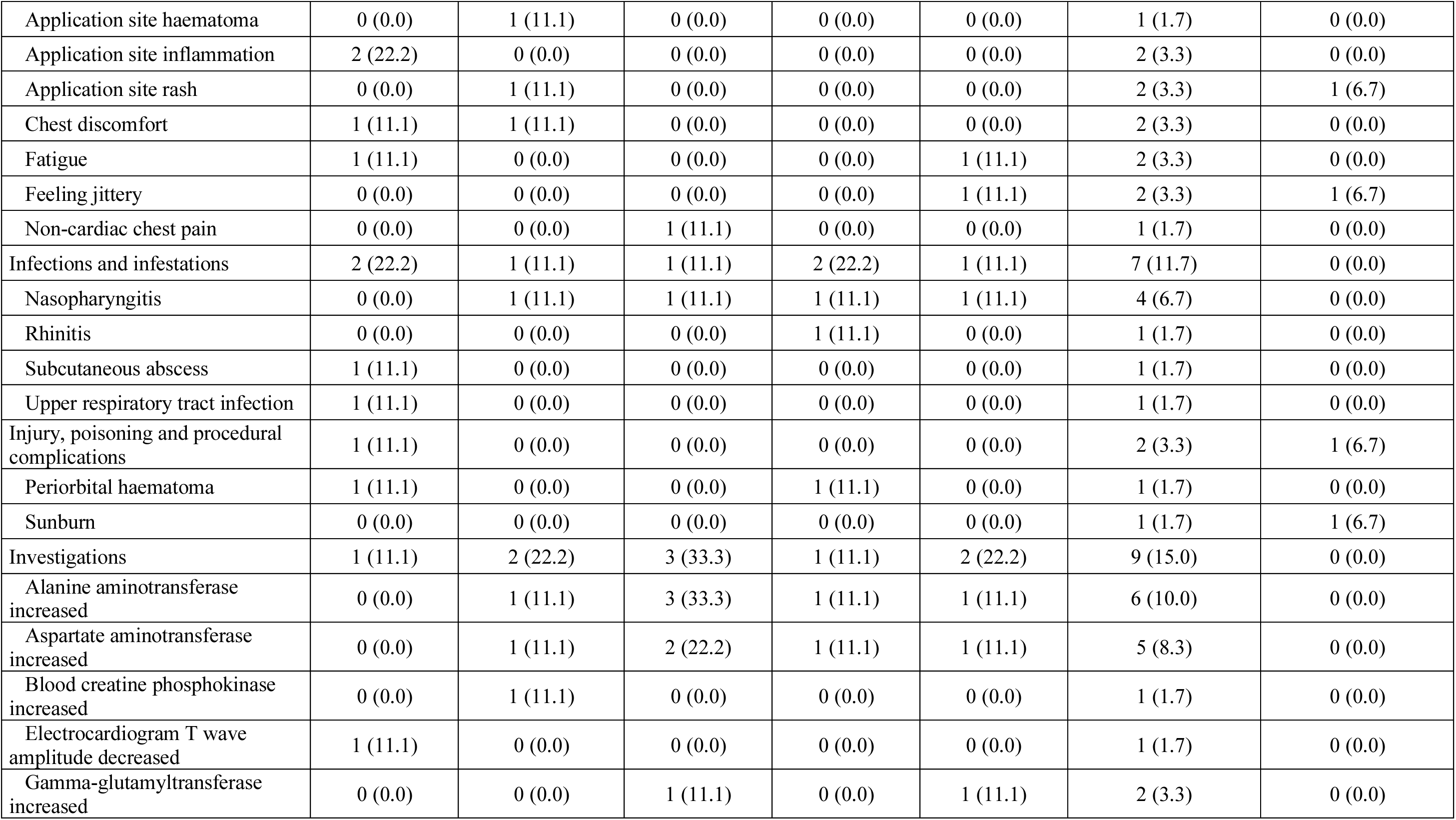
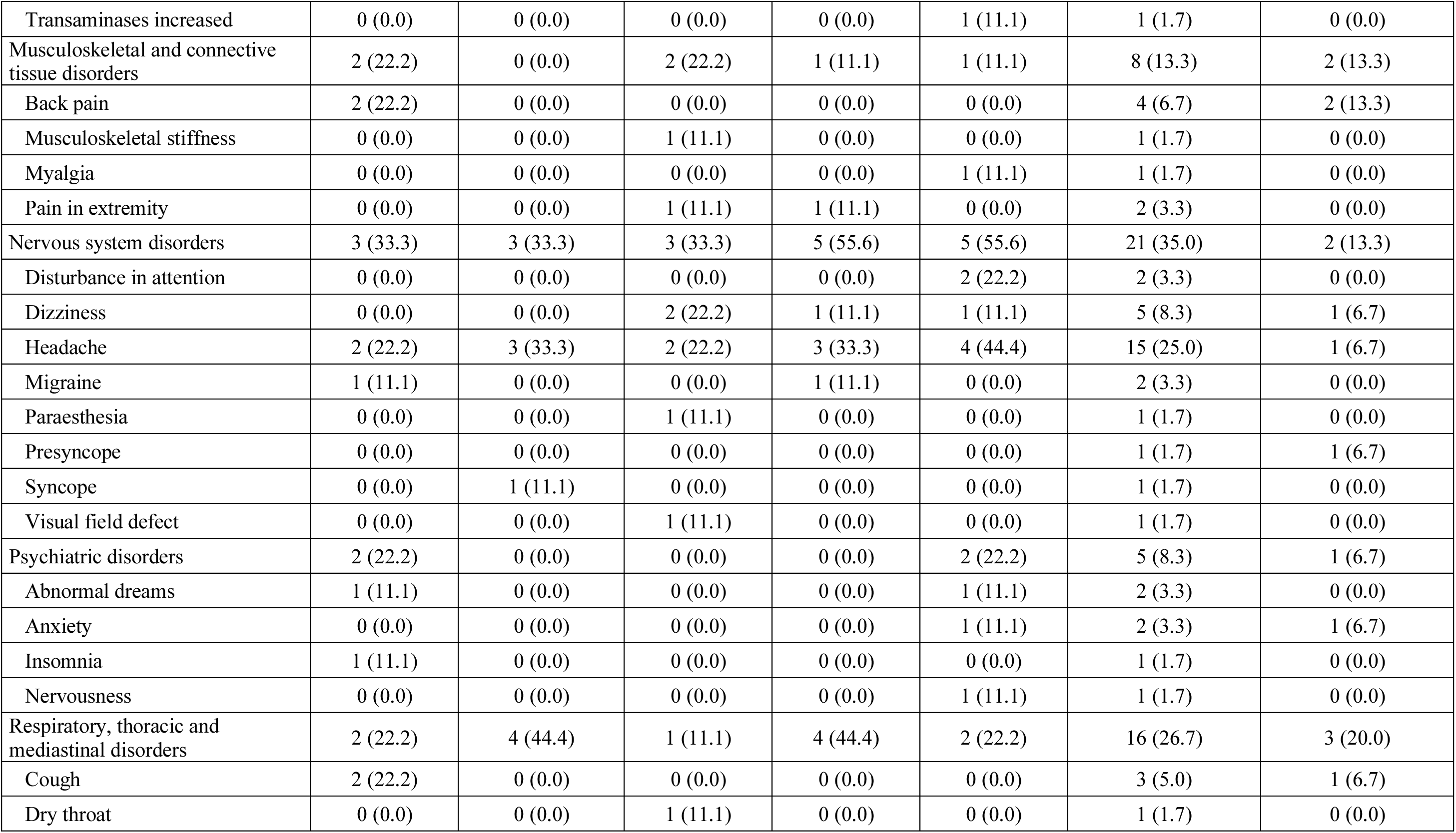
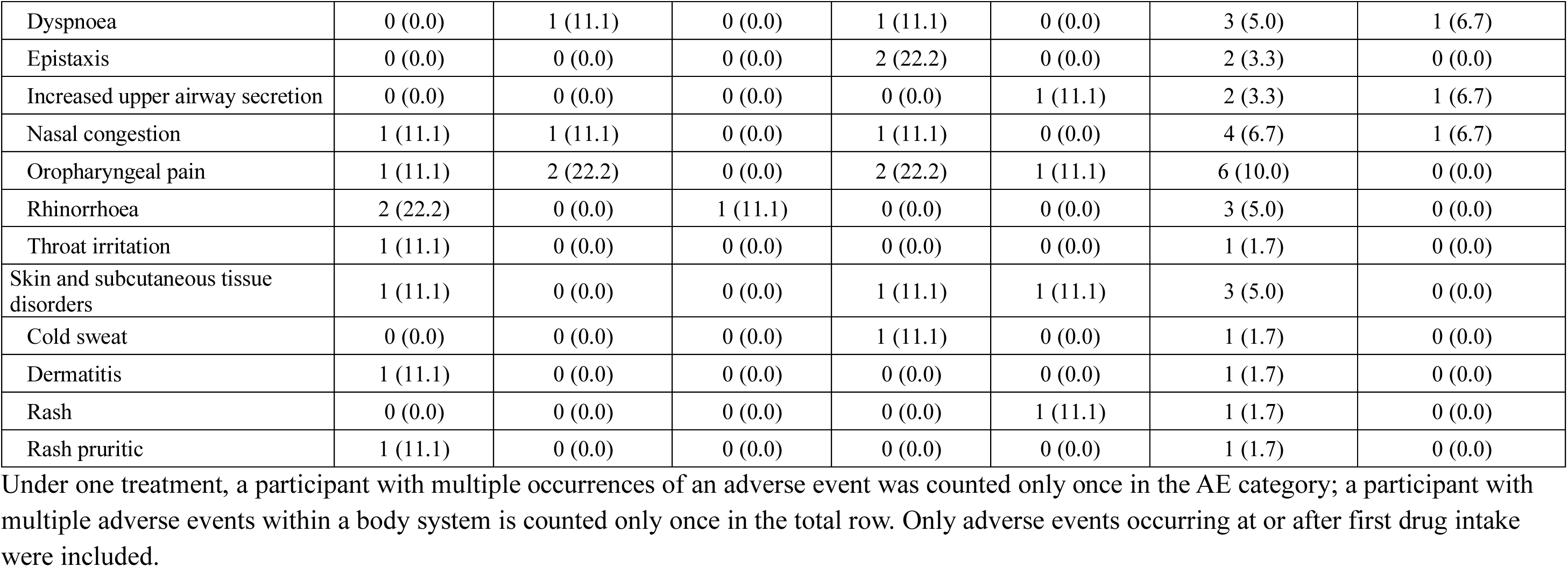
Adverse events overall and frequently affected system organ classes. **Table 4B**: Adverse events by body system and preferred term, safety analysis set

### Cardiac findings

Cardiac monitoring showed the expected negative chronotropic effect of mocravimod, with mean heart rates in active treatment groups decreasing to 50–56 bpm on Days 2–5 compared with a lowest mean of 57 bpm in placebo, and with the largest placebo-corrected reductions observed at 1.2–3.0 mg (10–11 bpm on Day 1, peaking at 14–15 bpm on Day 2); this effect attenuated over time, with smaller reductions by Day 14 and no meaningful difference from placebo by Day 28 in the 2.0 mg cohort, suggesting development of tolerance. Reductions in nadir heart rate (HR) during the first 24 hours were dose-dependent, with significant decreases observed from 0.6 mg onward and largest reductions (9–14 bpm) in the 1.2–3 mg groups, particularly on Day 2. The HR effect diminished over time, with smaller, less dose-dependent reductions by Day 5 and a return to near placebo levels by Day 14–28. Individual cases of mean heart rate <40 bpm occurred in the 0.6, 1.2, 2.0, and 3.0 mg groups, and marked sinus bradycardia was observed in one participant each in the 1.2 mg and 3.0 mg groups early after dosing. Flat T waves were noted in subject in the 0.3 mg arm on Day 14. The AE was graded mild in severity and suspected to be related to study drug. The mean QTcF intervals appeared to have no clear relationship to the dose administered or day of treatment. Mean Day 14 changes relative to the baseline in the QTcF for all cohorts were minor, ranging from −7.8 to 7.3 msec (the latter observed in the mocravimod 0.3 mg dose group) and the maximum QTcF interval in each treatment group was never above 450 msec any study day. Overall, there was no evidence of QT prolongation associated with mocravimod. Seven participants (2 placebo, 5 active) had brief, asymptomatic second-degree atrioventricular block (Mobitz I/Wenckebach) episodes, predominantly nocturnal in placebo and 0.6 mg recipients and mainly limited to the first 24 hours after dosing in active groups. One recurrent daytime Wenckebach event in the 3.0 mg group was reported as a mild, non-dose-limiting adverse event.

## Discussion and conclusion

The objectives of this Phase 1, multiple-ascending-dose, randomized, double-blind, placebo-controlled study were to assess the safety, tolerability, and PK of mocravimod in healthy volunteers. In sequential cohorts, mocravimod 0.3, 0.6, 1.2, 2 or 3 mg or placebo was administered once daily for 14 or 28 days.

PK of mocravimod and its active metabolite mocravimod-phosphate was close to steady-state conditions after 14 days of daily intake and at steady-state after 28 daily intakes. PK profiles of mocravimod-phosphate followed the same pattern as those of mocravimod, and both analytes circulated at similar concentrations, hence a mocravimod-phosphate / mocravimod molar ratio close to 1 and stable across doses. Furthermore, this ratio was similar between Day 1 and at steady-state suggesting that repeated dosing did not alter the metabolism of either analyte. This was also suggested by the approximate dose proportionality of exposure PK parameters AUC and C_max_. The estimated deviations from dose-proportionality are not considered clinically relevant. Accumulation of both analytes was predictable and exposure at steady state was of about 6 times the one after the first administration. This accumulation was similar across doses, consistent with the dose proportional behaviour of both the parent compound and its metabolite.

The lowest Day 1 level in the mean ALC for all mocravimod dosing groups occurred at approximately 4-6 hours post-dose. A dose dependent reduction of mean ALC was observed for all doses of mocravimod ≥0.6 mg. For the lowest dose, mocravimod 0.3 mg, no change was noted on Day 1 when compared to placebo. The apparent placebo effect on the ALC (29% reduction) may be a consequence of (1) post-dose values on the day of the first dose naturally lower than pre-dose values due to the circadian rhythm, and (2) the nature of the endpoint, i.e., the minimum of approximately 25 post-dose values which is likely smaller than the pre-dose value even in absence of a treatment effect. A comparison of the mean ALC at time zero (pre-dose) on Days 2, 4, 7, 10, 14, and in the morning of Days 21, 24, 28 and 35 to time zero on Day 1 for all dose groups, suggest that the initial post-dose mean ALC reduction on Day 1, remained suppressed throughout the 28 days of treatment. Maximal reduction rates occurred on Day 14 for all mocravimod dosing groups, except for cohort 1 (0.3 mg mocravimod) for which the lowest ALC was observed on Day 21. A dose dependent reduction of mean ALC was observed for all doses of mocravimod. The gradual return of absolute lymphocyte counts towards Day 1 pre-dose levels of the mean ALC after 14 days of daily mocravimod dosing was observed during the follow-up period. However full return to Day 1 pre-dose levels was not observed by the end-of-study (EOS) visit on Day 35 for any of the dose groups; a clear trend towards returning to Day 1 pre-dose levels was apparent and above the lower range of normal (i.e. > 1.0 109/L) by Day 28. Overall, the pharmacodynamic assessment demonstrated a predictable effect on ALC, in line with expectations. The mean reduction in ALC was dose dependent with the maximum effect appearing to approach plateau at the mocravimod 0.6 mg dose. A full return to Day 1 pre-dose levels was not observed by the end-of-study (EOS) visit on Day 35 for any of the dose groups; a clear trend towards returning to Day 1 pre-dose levels was apparent however and above the lower range of normal (i.e. >1.0 10^9^/L) by Day 28.

While the effects of S1P receptor modulators on the total leukocyte population have been extensively studied, less is known about their impact on specific leukocyte subsets in humans. Specifically, the effects of S1P receptor modulators on T- and B-cell subsets, such as on CD4+ and CD8+ T-cells, regulatory T-cells, and memory B-cells, as well as on innate cell populations have not been fully elucidated. According to a recent meta-analysis, treatment with any S1P receptor modulator is associated with an overall reduction in circulating CD4+ and CD8+ T lymphocytes. Decreases are also observed in naïve and central memory CD4+ and CD8+ T-cell subsets, although findings for effector memory T cells are inconsistent. Evidence regarding effects on other T-helper subsets (e.g., Th1 and Th17) is limited and therefore inconclusive, and changes in regulatory T cells beyond fingolimod have not been well characterized. Most studies report a consistent decline in B cells, including both naïve and memory populations, across indications. In contrast, no consistent treatment-related changes in peripheral blood monocyte or dendritic cell counts have been identified [9]. The leukocyte subset data suggest that the effect of mocravimod to dose dependently decrease the peripheral counts of different leukocyte subtypes was most pronounced on the B cells (CD19+B), the T cells (CD3+), the CD4+ and the CD8+ naïve cells (CD4N and CD8N) subsets in line with other S1P modulators. Monocytes, NK cells, CD4+ and CD8+ central memory (CD4CM and CD8CM) cells, dendritic cells, and plasma cells exhibited mixed effects (elevations and decreases from baseline) within a mocravimod dose level and between dose levels of mocravimod and no consistent treatment effect was evident. To assess a possible class effect, the cardiac effects of mocravimod were evaluated through 12-lead ECGs, 24-hour Holter, and 24-hour ABPM. No cardiac events were reported as serious adverse events or dose-limiting toxicity. The observed negative chronotropic effects were consistent with the known class effect of S1P receptor modulators, clinically benign, transient, and self-limiting, with tolerance developing over the 14–28 day dosing period. No QT prolongation was identified.

The possible pulmonary effects were assessed through pulmonary function tests. Other safety assessments included safety laboratory assessments to monitor for toxicity, CNS assessments in order to monitor for localized CNS accumulation due to chronic dosing, immunologic assessments to monitor for increased risk of infection possibly caused by the PD effects of mocravimod, and macular thickness assessment to monitor for visual disturbances and rare cases of macular oedema that have been reported with fingolimod in previous trials. The general lack of a relationship between doses and AEs as well as the lack of severity of the majority of the AEs, coupled with the generally transient nature of the AEs, suggest that mocravimod did not produce any definitive effect on the standard safety assessments in this study apart from the observed elevated liver function tests in some of the participants and the marked negative chronotropic effect on heart rate observed. Most of the participants with elevated liver function tests had a 2-3-fold transient increase in transaminase levels, which were either normalized or showed a clear trend towards normalization by the end of study. These changes in liver function tests are in line with previous observations from S1P receptor modulators. The cardiac effect (negative chronotropism), together with the significant ALC reductions were all characteristics of an S1P receptor modulator and did not result in any AEs.

The immunomodulatory properties of S1P receptor modulators have been exploited predominantly in autoimmune disorders; however, there is increasing clinical interest in their potential application in haematological malignancies and transplantation. In the setting of allo-HCT for AML, a central therapeutic challenge is the prevention of graft-versus-host disease (GvHD) while maintaining, or ideally enhancing, the graft-versus-leukaemia (GvL) effect [10, 11]. By limiting T-cell egress from secondary lymphoid organs, S1P receptor modulators reduce the trafficking of alloreactive T cells to GvHD target tissues, thereby offering a strategy to mitigate GvHD without compromising anti-leukaemic immune activity [7, 12]. Preclinical studies of mocravimod and other S1P receptor modulators have demonstrated that this approach can effectively “decouple” GvHD from GvL, resulting in improved survival in murine allo-HCT models. Consistent with these findings, early-phase clinical studies of mocravimod in patients undergoing allo-HCT for AML have reported encouraging safety and efficacy signals, including trends toward reduced relapse rates and improved overall survival compared with historical controls. The ongoing MO-TRANS Phase 3 trial is expected to provide definitive evidence supporting this therapeutic strategy [13]. If successful, mocravimod may represent a paradigm shift in the management of post-transplant relapse and GvHD, addressing a major unmet need in haematological oncology.

## Conclusion

Mocravimod was well tolerated, demonstrating a favourable safety profile. There were no findings or events that precluded escalation to each of the planned dose levels, treatment, SAEs or deaths. An expected, dose dependent reduction in ALC was observed with the maximum effect appearing to approach plateau at the mocravimod 0.6 mg once daily dosing. A full return to Day 1 pre-dose levels was not observed by the end of the study yet. The PK characteristics exhibited predictable accumulation, approximate dose-proportional exposure across the studied dose range and a stable metabolite to parent compound exposure ratio of about 1.

## Disclosures

## Data Availability

All data produced in the present study are available upon reasonable request to the authors.

## Acknowledgements

The authors wish to thank Dr. Margit Hemetsberger, Vienna, Austria, for editorial services, funded by Priothera SAS, Saint-Louis, France. This study was conducted by Novartis Pharma AG, Basel, Switzerland.

## Funding

Priothera SAS funded the open-access publication fees for this article.

## Conflicts of interest

All authors are employees of Priothera SAS.

## Author contributions

All authors have substantially contributed to the conception of the work; or the acquisition, analysis, or interpretation of data for the work. DH has drafted the work; DK and, JH reviewed it critically for important intellectual content. All authors have approved the final version of the work to be published and agree to be accountable for all aspects of the work in ensuring that questions related to the accuracy or integrity of any part of the work are appropriately investigated and resolved

## Notes

### Clinical Trial

EudraCT 2007-005608-42

### Author Declarations

Ethics committee of Edinburgh Independent Ethics Committee for Medical Research gave ethical approval for this work.

